# Shared genetics linking sociability with the brain’s default mode network

**DOI:** 10.1101/2024.05.24.24307883

**Authors:** Giuseppe Fanelli, Jamie Robinson, Chiara Fabbri, Janita Bralten, Nina Roth Mota, Martina Arenella, Emma Sprooten, Barbara Franke, Martien Kas, Till FM Andlauer, Alessandro Serretti

## Abstract

The brain’s default mode network (DMN) plays a role in social cognition, with altered DMN function being associated with social impairments across various neuropsychiatric disorders. In the present study, we examined the genetic relationship between sociability and DMN-related resting-state functional magnetic resonance imaging (rs-fMRI) traits.

To this end, we used genome-wide association summary statistics for sociability and 31 activity and 64 connectivity DMN-related rs-fMRI traits (N=34,691-342,461). First, we examined global and local genetic correlations between sociability and the rs-fMRI traits. Second, to assess putatively causal relationships between the traits, we conducted bi-directional Mendelian randomisation (MR) analyses. Finally, we prioritised genes influencing both sociability and rs-fMRI traits by combining three methods: gene-expression eQTL MR analyses, the CELLECT framework using single-nucleus RNA-seq data, and network propagation in the context of a protein-protein interaction network.

Significant local genetic correlations were found between sociability and two rs-fMRI traits, one representing spontaneous activity within the temporal cortex, the other representing connectivity between the frontal/cingulate and angular/temporal cortices. Sociability affected 12 rs-fMRI traits when allowing for weakly correlated genetic instruments. Combing all three methods for gene prioritisation, we defined 17 highly prioritised genes, with *DRD2* and *LINGO1* showing the most robust evidence across all analyses.

By integrating genetic and transcriptomics data, our gene prioritisation strategy may serve as a blueprint for future studies. The prioritised genes could be explored as potential biomarkers for social dysfunction in the context of neuropsychiatric disorders and as drug target genes.

**Highlights:**

- Genetic correlation identified between sociability and temporal cortex activity.
- Genetic correlation between sociability and frontal-angular/temporal connectivity.
- Sociability was causally linked to 12 MRI traits related to the default mode network.
- Seventeen prioritised genes were associated with both sociability and DMN traits.

## 1. Introduction

Sociability, defined as the inclination to seek or engage in social interactions, is a complex trait that manifests as a continuum within the general population (Caldwell, 2012, Reeb-Sutherland et al., 2012). Various physical and mental health-related outcomes are influenced by sociability (Cacioppo et al., 2015). Particularly important for the current study are the facts that social isolation is associated with mortality (Holt-Lunstad et al., 2015) and that social dysfunctions are relevant for neuropsychiatric conditions, including schizophrenia spectrum disorders, Alzheimer’s disease, autism spectrum disorder (ASD), and major depressive disorder (MDD) (Setien-Suero et al., 2022). Social dysfunctions constitute both a prodromal symptomatologic manifestation and a transdiagnostic negative prognostic factor (De Donatis et al., 2022, Oliva et al., 2022).

Several brain structures have been proposed as neural substrates of social behaviour (Porcelli et al., 2019), such as the praecuneus/posterior cingulate cortex and the medial-prefrontal and temporal regions (Porcelli, Van Der Wee, 2019). These brain areas are central nodes of the default mode network (DMN), an integrated network implicated in various higher-order functions including social cognitive processes (Buckner et al., 2008, Raichle et al., 2001). The DMN’s components serve discrete yet integrated functions. The rostromedial prefrontal cortex and posterior cingulate cortex are pivotal for processing socio-cognitive information with relevance to the self (Leech and Sharp, 2014, Smith et al., 2014). The medial temporal lobe, including the hippocampal formation, is essential for autobiographical memory processing, self-reflection, and the recollection of personal experiences (Andrews-Hanna et al., 2014, Spreng and Andrews-Hanna, 2015). The dorsomedial prefrontal cortex is integral to metacognitive functions, particularly those influenced by social context (Ferrari et al., 2016). Together with the temporoparietal junction, the dorsomedial prefrontal cortex enables mentalizing, i.e., the comprehension of mental states of other people (Andrews-Hanna, Smallwood, 2014, Spreng and Andrews-Hanna, 2015). The DMN’s role in self-referential cognition, theory of mind, the delineation of self from others, and autobiographical memory has been extensively documented (e.g., Andrews-Hanna et al. (2014), Spreng and Andrews-Hanna (2015)). The DMN has attracted considerable attention due to its pivotal role in mental functioning, as it integrates various cognitive processes such as self-reference, social cognition, episodic memory, and language (Menon, 2023) Recent studies have particularly highlighted the DMN’s relevance to social behaviours in the context of major psychoses, underscoring its function as a central integrative hub for coordinating cognitive activities related to both self-directed thought and responses to external stimuli (Fox et al., 2017, Mulligan and Bicknell, 2023, Saris et al., 2020). Our previous work has elucidated DMN functional connectivity alterations as potential transdiagnostic markers for social dysfunction (Saris et al., 2022), thereby underscoring the utility for an augmented genetic understanding of the network’s role in sociability (Gottesman and Gould, 2003).

Genetic factors influence both sociability and functional brain networks. The heritability of sociability-related behaviours, such as loneliness and social anxiety, was estimated at 48% (Boomsma et al., 2005, Stein et al., 2002), and the heritability estimate of DMN functional connectivity is 42% (Elliott et al., 2019, Glahn et al., 2010). Genome-wide association studies (GWAS) have identified single-nucleotide polymorphisms (SNPs) at 18 independent genomic loci to be associated with sociability (Bralten et al., 2021) and 45 genetic regions as associated with brain functional signatures (Zhao et al., 2022). Zhao and colleagues reported the SNP-based heritability (*h^2^_SNP_*) for resting state functional magnetic resonance imaging (rs-fMRI) node amplitude traits as 10.6-38.6% and for functional connectivity traits as 3-60%. While previous research demonstrated a phenotypic association between sociability and the DMN, the underlying genetic correlations, shared associated genes, or causal relationships have not yet been explored.

In the present study, we examined these genetic relationships by analysing patterns of global and local genetic correlations between sociability and DMN-related rs-fMRI traits and by exploring potential shared or causal genetic relationships between them. Global genetic correlation measures the overall genetic similarity between two traits across the entire genome, providing an estimate of the proportion of variance shared due to common genetic variants; this assessment includes pleiotropic effects, where a single genetic locus affects multiple traits (van Rheenen et al., 2019). Local genetic correlation examines restricted genomic regions to identify where traits show significant genetic overlaps, deviating from the genome-wide average (van Rheenen, Peyrot, 2019). Mendelian randomisation (MR), the third method used, leverages genetic variants as instrumental variables to infer causality between an exposure (e.g., sociability) and an outcome (e.g., DMN rs-fMRI traits) (Sanderson et al., 2022).

An improved understanding of the genetic underpinnings linking the DMN and sociability could support the identification of new biomarkers and treatment targets for social dysfunction in neuropsychiatric disorders and/or to the repurposing of existing drugs. Thus, the aims of the present study were, first, to characterise the genetic relationship between sociability and DMN-related rs-fMRI traits and, second, to prioritise genes associated with both types of traits. These prioritised genes can serve as a shortlist for future biomarker development and drug target discovery.

## 2. Methods

### 2.1. Input datasets

This study leveraged summary-level data from the largest available GWAS on sociability assessed from an aggregate score of UK Biobank self-report data (N=342,461; Bralten, Mota (2021)) and UK Biobank brain rs-fMRI traits based on independent component analysis (ICA) of resting-state neuroimaging data (N=34,691; Zhao, Li (2022)) (**Table S1**). Zhao et al. conducted rs-fMRI GWAS on 76 node amplitude traits measuring the intensity and temporal correlation of spontaneous BOLD activity within distinct network nodes, and between nodes, i.e., edges. These edges represent 1,695 measures of functional coactivity between pairs of network nodes and six global network dimensions of connectivity, derived through a dimensionality reduction process of the 1,695 pairwise connectivity traits (Zhao, Li, 2022). All the rs-fMRI traits mapping to the DMN, showing a significant *h^2^_SNP_*≥0.1, and with genome-wide significant loci (i.e., were available for download at https://zenodo.org/records/5775047), were selected for inclusion in our analyses. All utilized GWAS summary statistics included only individuals of European ancestry and used the reference genome build GRCh37/hg19.

### 2.2. Genetic overlap and causal relationships between sociability and rs-fMRI traits

We explored the genetic overlap between sociability and rs-fMRI traits related to the DMN using global and local genetic correlation analyses, as well as bi-directional MR analyses (**Fig. 1**). All rs-fMRI traits showing significant associations with sociability after correction for multiple testing in either correlation analyses or bi-directional MR analyses were selected for gene prioritisation.

**Figure 1.**
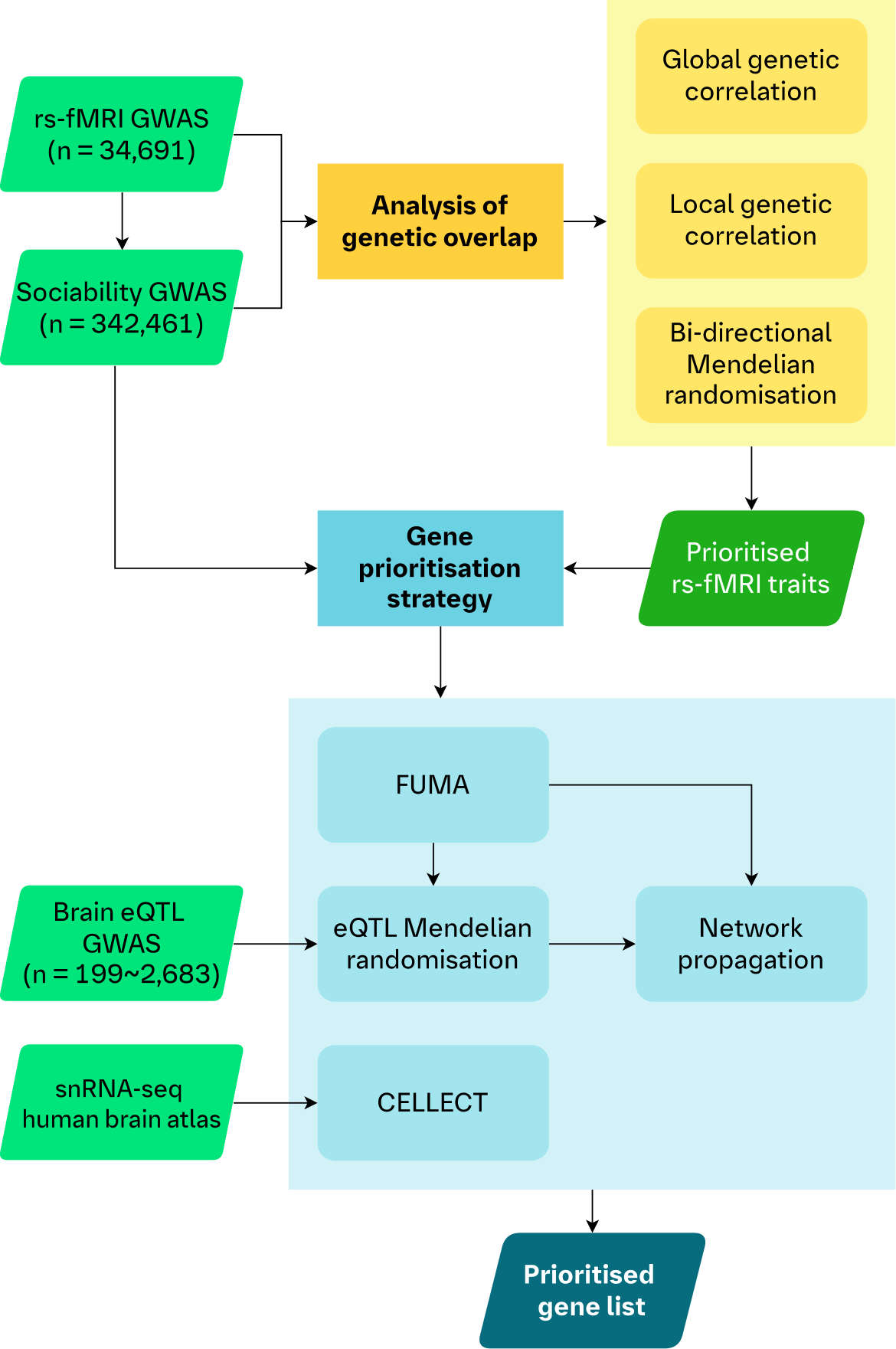
Analysis workflow. A schematic of the workflow of our analyses. We utilised genetic correlations and bi-directional MR to assess the genetic overlap between rs-fMRI traits and sociability to prioritise selected rs-fMRI traits for the downstream gene prioritisation strategy. First, the GWAS of the prioritised rs-fMRI traits and sociability were analysed using FUMA to map associated genetic regions to genes. We then leveraged eQTLs of gene expression in five brain tissues in an MR framework to provide further putative causal evidence for the mapped genes. Genes from these mapping steps were included in a TieDIE network propagation analysis using the underlying STRING protein-protein interaction network. Separately, we also integrated a human brain transcriptomics atlas (snRNA-seq data) in a CELLECT framework with the rs-fMRI and sociability GWAS. This step allowed us to identify genes whose increased expression are specific to cell types, in specific brain regions, for our traits of interest. Our final list of prioritised genes consisted of those genes which were identified by FUMA and showed at least nominal evidence in both the eQTL MR and CELLECT analyses, for both sociability and at least one rs-fMRI trait. Finally, we used the TieDIE network propagation scores to rank the list of prioritised genes.

#### 2.2.1. Global and local genetic correlation analyses

Bivariate Linkage Disequilibrium Score Regression (LDSC) analyses (Bulik-Sullivan et al. (2015); https://github.com/bulik/ldsc) were conducted to estimate the global genetic correlation (*r_g_*) between sociability and rs-fMRI traits using default LDSC parameters. LDSC is computationally robust even when sample overlap between GWASs occurs (Bulik-Sullivan, Finucane, 2015). Only the sociability and rs-fMRI trait pairs showing at least nominally significant (*p*<0.05) global, bivariate genetic correlations were further explored at the local genomic level.

Pairwise local genetic correlation analyses were conducted using LAVA (Local Analysis of [co]Variant Association), using standard parameters (Werme et al. (2022); https://github.com/josefin-werme/LAVA). In contrast to global correlation analyses, LAVA enables the identification of genomic regions that may be implicated in shared genetic aetiology and thus provides a fine-grained perspective on the genetic sharing between complex traits. Its analytical framework allows the investigation of genetic associations between traits under varying genetic causality scenarios without assuming a specific distribution of SNP effects (Werme, van der Sluis, 2022). In summary, we generated 2,495 semi-independent genomic regions of approximately 1 mega base-pair length, and only genomic regions with significant local *h^2^* (univariate *p*<1×10^-04^) for both phenotypes were analysed (Werme, van der Sluis, 2022). To account for overlapping samples among input GWASs, we supplied LAVA with intercepts derived from bivariate cross-trait LDSC analyses (Bulik-Sullivan, Finucane, 2015); these intercepts served as estimates of the sampling correlation between the datasets.

Because the rs-fMRI traits were not fully independent of each other, we applied the Benjamini-Hochberg correction for multiple testing, considering a maximum acceptable false discovery rate (FDR) of q=0.05 (Benjamini and Hochberg, 1995).

#### 2.2.2. Bi-directional Mendelian randomisation

We assessed the potential causal relationship between sociability and DMN-related rs-fMRI traits using bi-directional MR. Here, we parameterised two SNP instrument selection criteria following a paradigm employed previously to investigate a link between imaging-derived phenotypes and Alzheimer’s disease (Knutson et al., 2020): 1) a stringent analysis using only robust and independent instruments (*p*<5×10^-08^, clumping threshold *r^2^*<0.001), and 2) a lenient analysis using weakly-correlated instruments (*p*<5×10^-05^, clumping threshold *r^2^*<0.1). The rationale for this second approach was to increase the proportion of variance of the phenotype explained by the genetic variants and thus to increase statistical power (Knutson, Deng, 2020). Because these analyses were considered a complementary approach to genetic correlation analyses, sociability and all 95 DMN-related rs-fMRI traits were included. Multiple testing correction was applied using Bonferroni’s method based on the number of tested traits (significance threshold α=0.05/96=5.21×10^-04^).

#### 2.2.3. Mendelian randomisation statistical methods

We used the Wald ratio for exposures, which consisted of a single SNP instrument (overwhelmingly used in the eQTL MR, see below) or the inverse variance-weighted (IVW) method for exposures with multiple instruments (overwhelmingly used in the bi-directional MR, see above). However, we modified these methods to increase their utility for our use cases. First, we used the two-term Taylor series expansion of the Wald ratio to account for the error in both the instrument-exposure and instrument-outcome relationships. Second, we used an extended IVW method which allows for correlated instruments in the lenient bi-directional MR analyses (Burgess et al., 2016b). If a genetic exposure variant did not match the rsID of an outcome SNP, we searched for a proxy variant with a threshold of *r^2^*>0.8 (applies to all analyses).

To account for weak instrument bias, we specified that all instruments require an F-statistic of at least 10 (Bowden et al., 2016). As a sensitivity analysis for exposures with multiple instruments, we used an MR-Egger regression framework to assess the likelihood of the presence of horizontal pleiotropy. In the weakly-correlated instruments analysis, we used the extended method described by (Burgess, Dudbridge, 2016b). When analysing single genetic instruments, we examined evidence for reverse causation using Steiger filtering to assess whether the genetic variant explained more of the variance in the outcome than in the exposure (Hemani et al., 2017). Analyses were conducted using a custom implementation based on the TwoSampleMR R package (Hemani et al., 2018).

### 2.3. Identification of genes from GWAS loci

To identify genes associated with both sociability and the selected rs-fMRI traits, we conducted several complementary analyses providing causal, expression, and biological evidence (**Figure 1**). As a first step, we generated a list of potentially associated genes from the summary statistics of each GWAS using FUMA (Functional Mapping and Annotation of GWASs; Watanabe et al. (2017)), followed by providing causal evidence for the association of these genes using expression quantitative trait loci (eQTL)-based MR.

#### 2.3.1. FUMA-based gene mapping

We ran FUMA using default parameters for both sociability and the rs-fMRI traits prioritised in the genetic correlation and MR analyses to identify genes linked to the respective genome-wide significant loci. Only brain-derived tissue types were used for eQTL gene mapping. For prioritisation, genes had to meet any of the following criteria from the FUMA analyses: a) FDR-corrected MAGMA *p*<0.05; b) FDR-corrected eQTL mapping *p*<0.05; c) implicated in 3D chromatin mapping.

#### 2.3.2. Mendelian randomisation of gene expression

Using eQTL-based MR, the FUMA-prioritised genes were tested for evidence of a causal relationship with either sociability or the prioritised rs-fMRI traits. To this end, we estimated the putative causal effect of genetically proxied gene expression on genetic proxies of sociability and rs-fMRI traits. We utilised eQTLs from the MetaBrain resource, a meta-analysis of brain-derived eQTLs across five different tissues (basal ganglia, cerebellum, cortex, hippocampus, and spinal cord) (de Klein et al., 2023). We constructed *cis*-acting genetic instruments (as *trans*-acting eQTLs are more liable to pleiotropy) with a recursive *p*-value selection paradigm. We first searched for an eQTL for a gene at *p*<5×10^-08^ and, if no eQTL was found, reduced this threshold to *p*<5×10^-07^ and, finally, to *p*<5×10^-06^. If a gene was part of an eQTL with *p*<5×10^-08^, we did not include genetic variants with eQTL *p*-values in the range 5×10^-08^<*p<*5×10^-06^. Next, we ensured independence of SNPs by clumping at a threshold of *r^2^*<0.01. Our rationale for using a recursive *p*-value threshold was to include as many eQTLs, and thus genes, as possible in our gene prioritisation strategy. MR analyses were conducted for all identified eQTL genes. Multiple testing correction was applied using Bonferroni’s method based on the number of tested genes (significance threshold α=0.05/N_genes_). For further statistical details, see the Methods section *Mendelian randomisation statistical methods*.

### 2.4. Gene prioritisation strategy

To prioritise the identified genes further, we selected all genes that had at least nominal eQTL MR evidence for both sociability and at least one rs-fMRI trait, as well as at least nominal CELLECT evidence for both sociability and at least one rs-fMRI trait (*see below*). We ranked these genes using the TieDIE network propagation post-propagation score percentile (*see below*). We excluded genes mapping to the major histocompatibility complex region on chromosome 6 from our prioritised list due to the complex linkage disequilibrium structure in this region and the likelihood that the genetics-based analyses will not perform well there.

#### 2.4.1. Network propagation of genes

We used network propagation to identify additional genes affecting both sociability and the prioritised rs-fMRI traits in the context of protein-protein interaction (PPI) networks. We performed a two-seed node propagation in a tied diffusion through interacting events (TieDIE) framework to obtain genes which are close to seed genes from both sources (Paull et al., 2013). For this analysis, we selected all genes identified using FUMA that showed at least nominal eQTL MR evidence either for sociability or the rs-fMRI traits. These genes were used as seed nodes with binarised heats (i.e., if the gene was prioritised for the respective trait, the seed heat was ‘1’, otherwise it was ‘0’). As the underlying PPI network, we selected the full STRING database (Paull, Carlin, 2013) and removed interactions in the lowest quartile of all interaction scores (confidence score<0.309). We used the propagation algorithm implemented in the DiffuStats R package (Picart-Armada et al., 2018) and a jack-knife (i.e., leave-one-out) procedure for each network propagation analysis to ensure robustness of results against perturbations in the seed gene list. To aid the interpretation of results from this analysis, we grouped genes into percentiles based on the mean of their post-propagation scores across each of the jack-knife permutations.

#### 2.4.2. CELLECT cell type and gene identification

We used the CELL-type Expression-specific integration for Complex Traits (CELLECT) framework (Timshel et al., 2020) to integrate human single-nucleus RNA sequencing (snRNA-seq) datasets (Human Brain Cell Atlas v1.0, Siletti et al. (2023)) with the sociability and the prioritised rs-fMRI traits GWASs. For our analysis, we selected 22 dissections from this atlas covering representative brain regions related to the DMN, based on previous evidence (Ezama et al., 2021, Smallwood et al., 2021) (see **Table S2**). By leveraging partitioned LDSC and MAGMA, CELLECT identifies aetiologically important GWAS-associated cell types and genes which drive the cell type-phenotype association. Our correction for multiple testing thresholds were set as follows: for CELLECT identifying cell types, we used an FDR-corrected *p<*0.05; for CELLECT identifying genes, we used a Bonferroni threshold of *p<*1.01×10^-06^ (0.05/49,689 unique genes present in the MAGMA reference data).

## 3. Results

### 3.1. Genetic overlap and causal relationship

We analysed the genetic overlap of sociability with 31 activity (nodes) and 64 connectivity (edges) DMN-related rs-fMRI traits (**Table S1)**.

#### 3.1.1. Global and local genetic correlations of sociability with DMN-related activity and connectivity resting-state fMRI traits

After correcting for multiple testing, none of the global genetic correlations between sociability and rs-fMRI traits were significant (**Table S3**). The rs-fMRI phenotypes with the lowest nominal *p*-values were two amplitude traits, nodes 17 (temporal cortex) and 14 (cingulate/frontal cortex), and one connectivity measure, pair 15-45 (between insular/cingulate and frontal cortices), each showing a global genetic correlation of approximately *r_g_*=0.15 with sociability. Other nominally significant genetic correlations were found between sociability and three amplitude traits mapping to the insula, cingulate, parietal, and temporal cortex. These regions are involved in the DMN but extend to the motor, central executive, and salience networks. Nominally significant global genetic correlations with sociability were found for nine pairwise connectivity rs-fMRI traits and two whole brain functional connectivity measures involving the central executive, default mode, and salience networks of the triple-network model of psychopathology (Menon, 2011).

Local genetic correlation analyses were conducted using LAVA between sociability and the 16 DMN-related activity/connectivity rs-fMRI traits showing at least nominally significant global genetic correlations. A strong and significant local genetic correlation was observed between sociability and spontaneous activity in the temporal cortex (node 17) at chr18:34,153,298-36,056,932 (*r_g_*=0.66, *p*=2.1x10^-4^, *p_FDR_*=0.04) (**Table 1 and S4**). Among the rs-fMRI connectivity traits, a strong and significant local genetic correlation was identified at chr10:114,255,955-115,588,903 (*r_g_*=-0.70, *p*=1.7x10^-4^, *pFDR*=0.04), linking sociability with connectivity between the frontal/cingulate and angular/temporal cortex (edge 7-11) (**Table 1 and S4**). These areas are part of the DMN but extend to the limbic and central executive networks.

**Table 1.** Prioritised rs-fMRI traits. The rs-fMRI traits significantly locally genetically correlated with sociability (LAVA, see Table S4 for further details) or with significant MR evidence for sociability (Table S5), both after correction for multiple testing. Two traits with significant MR IVW *p*-values but evidence for pleiotropy are not shown. Effect size for LAVA: correlation *r^2^*; effect size for MR: IVW beta. The table shows uncorrected *p*-values. *Significant MR-Egger intercept *p*-value (indicating pleiotropy). The trait is shown nevertheless, because it also has significant local genetic correlation evidence through LAVA.

| rs-fMRI ID | Description | Location | Method | Effect size | P-value |
| --- | --- | --- | --- | --- | --- |
| **edge_pheno262** | Pair 7-11 | (Frontal\|Cingulate) & (Angular\|Temporal) | LAVA (local) | 0.49 | 1.67E-04 |
| **node_pheno17** | Node 17 | Temporal | LAVA (local) | 0.44 | 2.11E-04 |
| **edge_pheno262** | Pair 7-11 | (Frontal\|Cingulate) & (Angular\|Temporal) | MR (IVW lenient) | -0.34 | 9.75E-12 |
| **edge_pheno1142** | Pair 29-44 | Parietal & Frontal_Inf | MR (IVW lenient) | 0.24 | 1.14E-06 |
| **edge_pheno816** | Pair 11-36 | (Angular\|Temporal) & Precuneus | MR (IVW lenient) | -0.17 | 3.15E-06 |
| **edge_pheno491** | Pair 5-25 | (Precuneus\|Angular\|Cingulate) & (Frontal\|Precentral) | MR (IVW lenient) | -0.17 | 4.91E-06 |
| **edge_pheno1698** | ICA 3 | Global measure | MR (IVW lenient) | -0.22 | 1.80E-05 |
| **edge_pheno1205** | Pair 5-46 | (Precuneus\|Angular\|Cingulate) & Temporal | MR (IVW lenient) | 0.18 | 2.18E-05 |
| **edge_pheno253** | Pair 7-10 | (Frontal\|Cingulate) & (OccipitalPrecuneus) | MR (IVW lenient) | 0.16 | 2.92E-05 |
| **node_pheno17** | Node 17 | Temporal | MR (IVW lenient) | 0.18 | 1.01E-04* |
| **edge_pheno249** | Pair 3-10 | (Lingual\|Fusiform) & (OccipitalPrecuneus) | MR (IVW lenient) | -0.16 | 1.35E-04 |
| **edge_pheno593** | Pair 5-29 | (Precuneus\|Angular\|Cingulate) & Parietal | MR (IVW lenient) | -0.15 | 2.61E-04 |
| **edge_pheno1359** | Pair 21-49 | Frontal_Sup & (Temporal_Mid\|Angular) | MR (IVW lenient) | 0.14 | 3.70E-04 |
| **edge_pheno286** | Pair 10-13 | (OccipitalPrecuneus) & Frontal_Sup | MR (IVW lenient) | 0.14 | 4.63E-04 |

#### 3.1.2. Assessment of putatively causal effects using bi-directional MR

In the stringent bi-directional MR analysis, no results remained significant after Bonferroni correction for multiple testing. The putatively causal genetically proxied relationship showing the lowest nominal *p*-value (*p*=1.42×10^-03^) was observed for sociability on the rs-fMRI edge 31-48 (connectivity between frontal and temporal cortex) (**Table S5**, MR method “Inverse variance weighted”).

In the lenient bi-directional MR analysis, we found evidence for a genetically proxied causal effect of sociability on 14 rs-fMRI traits, 13 of which were connectivity measures (**Table 1 and S5**, MR method “Inverse variance weighted correlated”). The rs-fMRI trait showing the most robust MR evidence for being causally affected by sociability was edge 7-11 (*p*=9.75×10^-12^, which also showed evidence for local genetic correlation (*p*=1.7×10^-4^, **Table S4**)). Node 17, which also exhibited significant local genetic correlation with sociability, was the only activity rs-fMRI trait significant after correction for multiple testing (*p*=1.01×10^-04^). However, we found evidence for horizontal pleiotropy (intercept *p*=6.94x10^-04^) in the MR-Egger analysis for this node. Because this node was independently prioritised using local genetic correlation, it was retained in the gene prioritisation analyses. Two additional edges (44-49 and 10-36) that showed evidence for pleiotropy as indicated by MR-Egger (**Table S5**), were excluded from further analyses, leaving 12 prioritised rs-fMRI traits.

### 3.2. Gene prioritisation

#### 3.2.1. Mapping GWAS loci to genes using FUMA and MR

After establishing that sociability and 12 DMN-related rs-fMRI traits were either locally correlated (LAVA) or putatively causally related (bi-directional MR), we prioritised genes potentially affecting both phenotypes (**Figure 1**). Using FUMA locus-to-gene analyses for each GWAS, we mapped 83 genes for sociability and 172 unique genes across the 12 rs-fMRI traits (**Table S6**). The median number of genes identified per rs-fMRI trait was 4.5, ranging from zero (edge 11-36) to 92 (edge ICA 3). Among these genes, eQTL-based MR analyses prioritised nine unique genes as significantly associated with sociability (*p*<0.05/83) and 32 unique genes significantly associated with rs-fMRI traits (*p*<0.05/172) (**Table S7**).

#### 3.2.2. Prioritised of cell types and genes associated with DMN and sociability

CELLECT-based cell type analyses reached the lowest *p*-values for deep-layer intra-telencephalic neurons across the different cortex dissections (**Table S8-S9**). In addition to 54 seed genes (selected using FUMA and eQTL-MR), 139 novel genes were in the top percentile of network propagation results (**Table S10**).

When combining gene prioritisation methods, 43 genes showed nominal eQTL-based MR and CELLECT evidence for both sociability and at least one rs-fMRI trait (**Table S11**). Most of these genes (31 of 43) had eQTLs derived in the cortex, which is likely due to the larger sample size for this tissue compared to the other brain regions. Furthermore, ten of the 43 genes were in the top decile of network propagation results, indicating they acted closely in PPI networks to the seed genes of both sociability and rs-fMRI traits (**Table S11**). Only six of these 43 prioritised genes were also identified using FUMA (**Table 2**). Of these, only *DRD2* and *LINGO2* fulfilled all prioritisation criteria, showed eQTL MR evidence significant after correction for multiple testing in any of the five tested brain regions, and were in the top percentile of network propagation.

**Table 2.** Top prioritised genes. From the list of 43 prioritised genes (Table S11), the 17 genes prioritised by FUMA, showing a significant eQTL MR *p*-value after correction for multiple testing with either sociability or any of the rs-fMRI traits, exhibiting a significant CELLECT MAGMA *p*-value after correction for multiple testing, or being in the top decile of network propagation. Significant *p*-values after correction for multiple testing are shown in bold font.

| Gene | FUMA Prioritised |  | eQTL MR |  | CELLECT |  | TieDIE |
| --- | --- | --- | --- | --- | --- | --- | --- |
| Gene Symbol | Prioritised Sociability | Prioritised rs-fMRI | Min P for Sociability | Min P for rs-fMRI | Min P Sociability | Min P rs-fMRI | Score Percentile |
| *DRD2* | Yes | No | **1.36E-04** | 4.80E-03 | **5.09E-10** | 4.67E-06 | **100** |
| *LINGO1* | Yes | No | **6.00E-04** | 4.37E-02 | **9.74E-08** | 2.15E-02 | **100** |
| *ELAVL2* | Yes | No | 9.15E-04 | 3.15E-02 | **3.15E-10** | 5.98E-05 | **100** |
| *CTNND1* | Yes | No | 7.08E-03 | 1.77E-02 | **7.71E-08** | 2.92E-02 | **100** |
| *YJEFN3* | No | No | **6.49E-05** | 2.04E-03 | 4.18E-04 | 9.65E-03 | **98** |
| *AMZ1* | No | No | 3.89E-03 | 2.71E-03 | 2.68E-03 | 9.56E-04 | **98** |
| *LST1* | No | No | 4.00E-02 | 4.32E-03 | 2.91E-04 | 1.83E-02 | **97** |
| *ANKK1* | No | No | 1.62E-02 | 2.54E-02 | **3.98E-08** | 7.60E-05 | **95** |
| *MIXL1* | No | No | 3.69E-03 | 2.25E-02 | 1.11E-04 | 2.73E-02 | **93** |
| *NOTCH4* | No | No | 3.99E-02 | 2.62E-02 | 2.84E-03 | 2.12E-02 | **90** |
| *ZIC1* | No | Yes | 2.42E-02 | 3.43E-02 | 9.63E-03 | 3.36E-06 | 88 |
| *KPNA2* | No | Yes | **1.78E-04** | **1.02E-05** | 6.69E-05 | **1.37E-08** | 76 |
| *MDGA1* | No | No | **2.23E-04** | 1.93E-02 | 9.75E-03 | 8.57E-03 | 47 |
| *FRAT1* | No | No | **2.64E-04** | 1.76E-02 | 2.93E-03 | 1.21E-02 | 42 |
| *SYBU* | No | No | **2.68E-05** | **2.29E-04** | 2.75E-06 | 1.87E-04 | 35 |
| *EMB* | No | No | **7.66E-05** | **8.28E-05** | 1.25E-03 | 3.11E-03 | 30 |
| *SYCE1L* | No | No | **5.29E-04** | 2.37E-02 | 7.60E-04 | 2.88E-02 | 12 |

Of the 43 prioritised genes, 11 were associated with anxiety and ten with depressive symptoms (Thorp et al., 2021), ten with various neuroticism items (Nagel et al., 2018), and seven with risk-taking behaviour (Thorp, Campos, 2021). According to the Open Targets Platform (https://www.opentargets.org), 11 of the 44 genes show an indication of being druggable, *i.e.*, fulfil any criteria for small molecule tractability.

## 4. Discussion

In the present study, we investigated the genetic relationship between sociability and default mode network (DMN)-related resting-state functional magnetic resonance imaging (rs-fMRI) traits, unveiling novel insights on the neural and genetic underpinnings of social behaviour. We leveraged robust analytical frameworks including genome-wide global and local genetic correlation analyses, bi-directional Mendelian randomisation (MR), and a comprehensive gene prioritisation strategy.

Significant local genetic correlations were found between sociability and specific DMN-related rs-fMRI activity and connectivity traits, suggesting shared pathophysiology, with subsequent analyses revealing putatively causal relationships between sociability and 12 rs-fMRI traits. Our gene prioritisation approach, integrating eQTL-based MR, sn-RNAseq, and network propagation analyses, highlighted 43 genes with evidence supporting their influence on both sociability and DMN-related rs-fMRI traits, among which *DRD2* and *LINGO2* were most consistently implicated and emerged as key targets of interest.

Our study is the first to find significant genetic correlations between sociability and resting-state neural activity mapping to the DMN, within the temporal, cingulate, and frontal cortices. The temporal cortex, particularly the superior temporal sulcus, is involved in the perception of social interactions. This lobe’s involvement extends across various cognitive domains essential for social behaviour, including facial recognition, communication, and emotion processing (Deen et al., 2015). The cingulate cortex, intersecting with the DMN (through the posterior cingulate cortex) but also with the salience network (SN), is essential for detecting and orienting attention toward salient stimuli and for cognitive control modulation on a trial-by-trial basis (Bartoli et al., 2018). In particular, the anterior cingulate cortex (ACC) processes social information and detects subjectively rewarding opportunities in social evaluation by assessing others’ behaviours and motivation (Apps et al., 2016, Rigney et al., 2018). The frontal cortex, especially the prefrontal cortex, underlies higher cognitive functions and social adaptive skills which are indispensable for complex social interactions (Firat, 2019). The medial prefrontal cortex, a component of the DMN, is implicated in early social cognition and enables us to behave adequately across specific social contexts https://doi.org/10.3389/fnhum.2013.00340 (Grossmann, 2013).

The three brain cortical regions, overlapping with the DMN, are also functionally involved in the motor network (MN), central executive network (CEN), and SN, highlighting the importance of inter-network functionality for social behaviour. The motor network’s role in social cognition, though less explored, includes the comprehension of movements and intentions by others, a core component of social understanding (Henschke and Pakan, 2023). The CEN, primarily involving the dorsolateral prefrontal cortex and posterior parietal cortex, is associated with high-level cognitive functions, including working memory, cognitive control, and decision-making (Menon, 2011). The SN, encompassing the ACC and insula, plays a role in detecting and orienting attention toward salient stimuli, and in switching between the DMN and CEN (Shaw et al., 2021). The involvement of the CEN and SN, in particular, may reflect the relevance of cognitive control and the identification of salient stimuli to social cognition (Nazlidou et al., 2015, Rijpma et al., 2021). Resting-state activity within these regions and their connectivity can influence social behaviour. For instance, aberrant activity and connectivity within these networks have been linked to altered social behaviour and cognitive functions observed in conditions such as ASD and MDD (Blume et al., 2023, Rolls et al., 2020).

Our local genetic correlation analyses revealed that sociability is genetically linked to brain connectivity between the frontal/cingulate and angular/temporal cortex. These findings extend beyond the DMN and implicate areas within the limbic system and the CEN. The connectivity between these areas might facilitate switching between self-referential thought (typical of DMN function) and external attention-demanding tasks, as well as the integration of emotional processing, thereby enabling effective social interaction. Interestingly, functional connectivity within and between the frontal and temporal lobes is modulated by social context, and enhanced when more difficult social inferences are made (Ainsworth et al., 2021).

Only two of the 95 DMN-related rs-fMRI traits analysed were significantly correlated with sociability in the LAVA analysis and showed significant MR evidence. However, none of the global correlation and stringent MR analyses were significant after correction for multiple testing. Given the relatively high *h^2^_SNP_* of neuroimaging traits, estimated to be between 20-40% for fMRI traits (Adhikari et al., 2018, Elliott et al., 2018), this low number of significant results might appear surprising. However, high *h^2^_SNP_* does not necessarily directly confer high statistical power for detecting causal loci (see Fan et al. (2018)). In general, the complex and unique genetic architecture of imaging traits makes correlational analyses difficult (Toro et al., 2015). In fact, several seemingly well-powered studies have failed to detect significant genetic correlations between MRI-derived phenotypes and psychiatric traits (see Andlauer et al. (2021) and the discussion therein). Furthermore, measured rs-fMRI traits are somewhat distant from their molecular effectors because they integrate various genetic and environmental influences plus technical batches and other sources of heterogeneity (Andlauer, Muhleisen (2021). In our opinion, it is justified under these conditions to increase the search space for the causal MR analysis. It has been demonstrated that including multiple correlated instruments in an MR framework increases statistical discovery power particularly for imaging-derived traits where few loci reach genome-wide significance (Knutson and Pan, 2021). Thus, we adopted this approach also for the present study and thereby found strong evidence that genetic proxies of sociability putatively influence various DMN-related rs-fMRI measures.

Our gene prioritisation strategy used triangulation – a key practice in aetiological epidemiology – whereby more reliable results can be obtained when integrating analyses from different datasets and methods with orthogonal sources of bias (Lawlor et al., 2016). Such an approach can be especially useful when evidence from one method alone is not considered as robust enough. We employed triangulation to integrate diverse evidence for genes potentially affecting both sociability and DMN-related rs-fMRI measures. The validity of this approach is underscored by many highly interesting examples of genes prioritised in the present study.

The fact that the gene coding for the dopamine receptor D2 (*DRD2*) was highly prioritised using our approach can be considered as a positive control. DRD2 is the target of anti-psychotic medications and consistently appears as a significant locus across GWAS for psychiatric disorders and symptoms, including major depression (Meng et al., 2024), depressive symptoms (Nagel, Watanabe, 2018), anhedonia (Ward et al., 2019), suicide (Kimbrel et al., 2022), and addiction (Kimbrel, Ashley-Koch, 2022). The neighbouring gene, coding for the protein kinase ANKK1, was also in the top network propagation decile and is associated with, among others, major depression (Meng, Navoly, 2024), bipolar disorder (Bipolar Disorder Working Group of the Psychiatric Genomics Consortium et al., 2023), and addiction (Hatoum et al., 2023). Notably, *ANKK1* was, unlike *DRD2*, not a network prioritisation seed gene and hence identified by network propagation. Several other interesting genes already studied in the context of brain development and function and with previous human genetics evidence were identified in the present study. One of these genes is *LINGO1*, which was, next to *DRD2*, the second gene identified across all prioritisation methods. It is a regulator of large conductance Ca^2+^-activated potassium channels (Dudem et al., 2020) and is associated with depressive symptoms (Dudem, Large, 2020) and addiction (Liu et al., 2019, Saunders et al., 2022). It was also reported to be associated with cognitive function in schizophrenia (Andrews et al., 2023, Fernandez-Enright et al., 2014). This gene has been suggested as a drug target (Andrews and Fernandez-Enright, 2015) but has low potential for small-molecule tractability (https://platform.opentargets.org/). Another example of an interesting prioritised gene is the regulator of protein translation *ELAVL2*, with a potential role in neurodevelopment (Mulligan and Bicknell, 2023). The *ELAVL2* gene is also associated with, among other psychiatric traits, major depression (Wainberg et al., 2022) and cognitive function (Davies et al., 2018).

### 4.1. Strengths and limitations

To the best of our knowledge, the present study was the first to find a genetic link between sociability and the DMN, expanding previous evidence of phenotypic associations (Li, Mai, 2014, Saris, Aghajani, 2022). To this end, we leveraged the largest GWAS summary statistics available for both sociability and the rs-fMRI traits. Another strength of our study was the integration of various analysis methods, with the aim of complementing and triangulating evidence. To overcome issues inherent to imaging genetics, we used previously validated instruments to increase the statistical power of our causal inference methods. By integrating genetic and transcriptomics data as well as statistical genetic and network-based methodologies, our gene prioritisation strategy may serve as a blueprint for future studies.

There are also limitations to our study. The statistical power of our genetic correlation analyses was likely reduced by the complex genetic structure of both sociability and the rs-fMRI traits, as e.g. evidenced by the low *h^2^* of the sociability GWAS (∼6%). Secondly, the CELLECT framework has the inherent limitation to only identify genes whose increased expression affects a given trait, but not genes whose decreased expression affects either sociability or the rs-fMRI traits. In the MR analyses, we could not rule out the presence of horizontal pleiotropy, i.e., the observed effect being mediated through an unrelated pathway. However, we used methods like MR-Egger regression to assess the likelihood of pleiotropy taking place and filtered the results accordingly. To this end, we employed an expansion of the standard MR-Egger method to account for weakly correlated instruments (Burgess, Dudbridge, 2016b). Finally, the GWAS of sociability and rs-fMRI traits used in the bi- directional MR analysis were derived using participants in UK Biobank, and sample overlap is a potential concern in two-sample MR (so-called “winner’s curse bias”). However, previous papers on this subject have indicated that, in such a case, weak-instrument bias is amplified, and the inflation of the false discovery rate is moderate to low (Sadreev et al., 2021). To partially address and mitigate the effects of weak-instrument bias, we corrected for multiple testing using a more stringent threshold (Bonferroni’s threshold instead of FDR), and we also ensured that all instruments were subject to an F-statistic threshold. Nevertheless, we caution that this measure is likely not sufficient to alleviate any potential bias in the causal effect estimates (Burgess et al., 2016a).

### 4.2. Conclusions

In conclusion, our study identified genetic factors common to sociability and functional measures of DMN activity and connectivity. The significant rs-fMRI nodes and edges primarily cover the temporal, cingulate, and frontal cortex – brain regions integral to the DMN. Intriguingly, these regions also extend to the motor, central executive, and salience networks. The present study paves the way for further exploration into the clinical translation of these findings, holding the potential to inform biomarker development and design of novel therapeutic strategies for neuropsychiatric disorders featuring social impairment.

## Data Availability

All data produced in the present work are contained in the manuscript and/or supplementary materials.

## Acknowledgements

The PRISM2 project (https://prism2-project.eu/en/prism-study/) leading to this application has received funding from the Innovative Medicines Initiative 2 Joint Undertaking under grant agreement Number 101034377. The work also was supported by funding from the European Community’s Horizon 2020 Programme (H2020/2014 – 2020) under grant agreement Number 847879 (PRIME) and the National Institute of Mental Health of the National Institutes of Health under Award Number R01MH124851. This publication reflects only the authors’ views, neither IMI JU, EFPIA, the European Commission, nor the National Institutes of Health are liable for any use that may be made of the information contained therein.

